# Low-Impact Social Distancing Interventions to Mitigate Local Epidemics of SARS-CoV-2

**DOI:** 10.1101/2020.06.30.20143735

**Authors:** Michael L. Jackson

## Abstract

**Background:** After many jurisdictions have implemented intensive social distancing to suppress SARS-CoV-2 transmission, the challenge now is to mitigate the ongoing COVID-19 epidemic without overburdening economic and social activities. This report explores “low-impact” interventions to mitigate SARS-CoV-2 with a minimum of social and economic disruption.

**Methods:** An agent-based model simulated the population of King County, Washington, with agents that interact in homes, schools, workplaces, and other community sites. SARS-CoV-2 transmission probabilities were estimated by fitting simulated to observed hospital admissions from February – May 2020. Interventions considered were (a) encouraging telecommuting; (b) reducing contacts to seniors and nursing home residents; (c) modest reductions to contacts outside of the home; (d) encouraging self-isolation of persons with COVID-19 symptoms; (e) rapid testing and household quarantining.

**Results:** Individual interventions are not expected to have a large impact on COVID-19 hospitalizations. No intervention reduced COVID-19 hospitalizations by more than 12.7% (95% confidence interval [CI], 12.0% to 13.3%). Removing all interventions would result in nearly 42,000 COVID- 19 hospitalizations between June 2020 and January 2021, with peak hospital occupancy exceeding available beds 6-fold. Combining the interventions is predicted to reduce total hospitalizations by 48% (95% CI, 47-49%), with peak COVID-19 hospital occupancy of 70% of total beds. Targeted school closures can further reduce the peak occupancy.

**Conclusions:** Combining low-impact interventions may mitigate the course of the COVID-19 epidemic, keeping hospital burden within the capacity of the healthcare system. Under this approach SARS-CoV-2 can spread through the community, moving toward herd immunity, while minimizing social and economic disruption.

## Background

In the absence of an effective and widely available vaccine against SARS-CoV-2, social distancing measures are the primary interventions available for mitigating the severity of the COVID-19 pandemic. During the initial pandemic wave, many jurisdictions around the world implemented intensive social distancing measures, such as forbidding large gatherings, closing schools and businesses, or issuing shelter-in-place orders.^1,2^ These measures have, for the most part, successfully suppressed transmission of SARS-CoV-2 by lowering the effective reproductive number of the virus.^2-4^

While effective at reducing SARS-CoV-2 transmission, these social distancing measures have incurred high economic costs^5^ and are now being relaxed in many jurisdictions. The challenge is to identify economically and socially sustainable interventions that can keep COVID-19 incidence low enough to be managed with existing hospital capacity. This report presents findings from an agent-based model of SARS-CoV-2 transmission that can help guide decisions about mitigating the impact of COVID-19 during this re-opening. The specific focus is identifying “low-impact” interventions – interventions which cause a minimum of disruption to economic and social activity – that can be combined to mitigate the ongoing COVID-19 epidemic.

## Results

The agent-based model tracks individual members of a simulated population modeled after King County, Washington, with demographic and household composition based on the 2018 American Community Survey.^6^ The per-contact probability of transmitting SARS-CoV-2 in homes and in non-home settings was estimated by fitting simulated daily COVID-19 hospitalizations to hospitalizations in King County from 28 February – 27 May 2020, in the presence of social distancing interventions as actually implemented in King County.^7^ These transmission probabilities correspond to an estimated basic reproductive number was 2.7 (95% CI, 2.4 – 3.1).

The model was able to reproduce key features of COVID-19 incidence in King County, including a rapid rise to a peak of approximately 40 admissions per day in late March and early April, followed by a steady decline through the end of May (Figure 1a). The model also generally captured the pattern of COVID-19 hospitalizations by age, with very few hospitalizations among children and adults aged <20 years and the greatest number among adults aged 60-79 years (Figure 1b).

**Figure 1:**
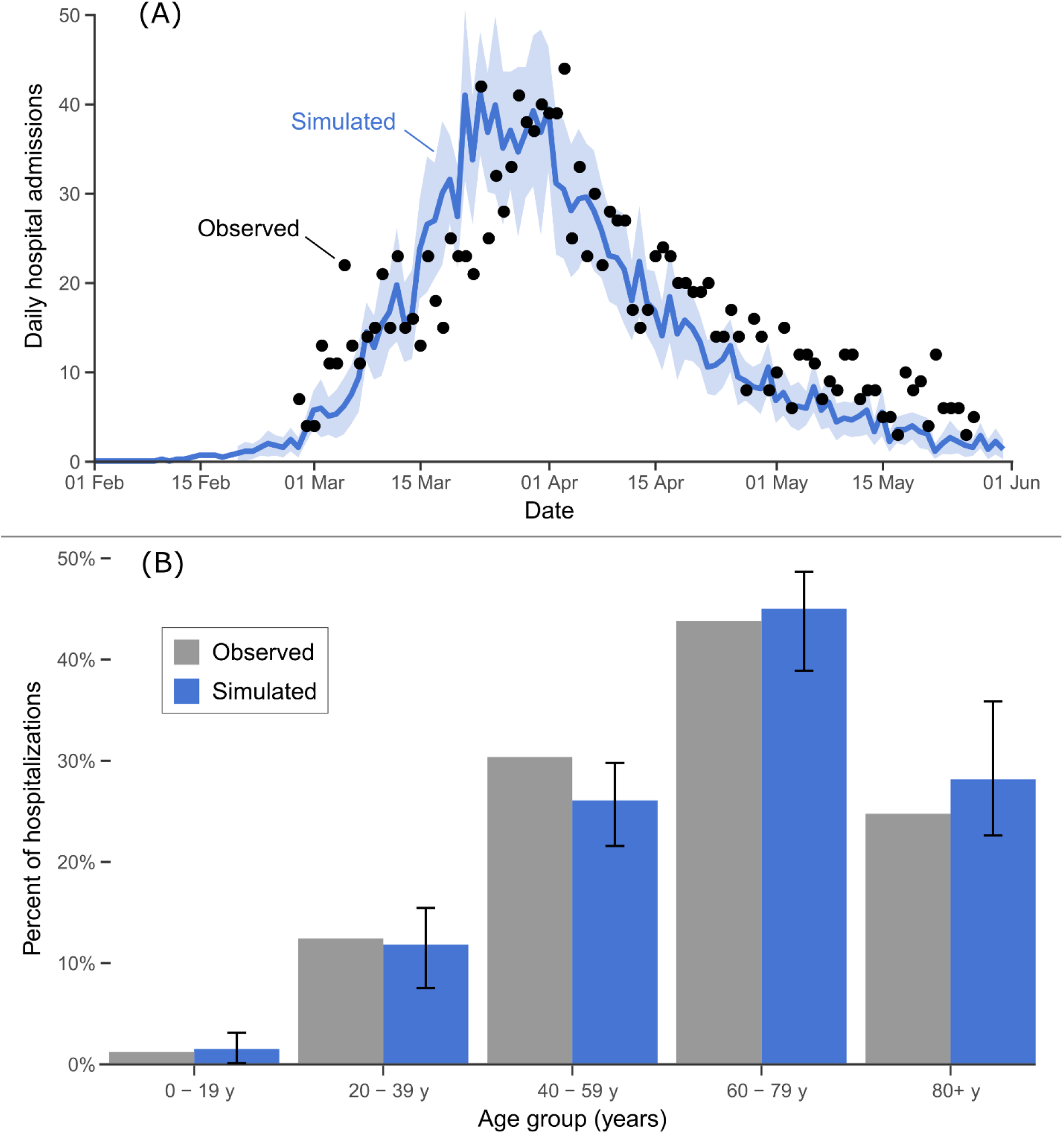
Observed vs. simulated count of hospital admissions for COVID-19 – King County, February – May 2020. Panel A, admissions by date; Panel B, admissions by age

Next, several interventions were considered. These were aimed at reducing SARS-CoV-2 transmission without having a high impact on economic and social activity in King County. The modeled interventions were:

- **Voluntary work-from-home**. Under this intervention, employers are encouraged to use telecommuting or other remote work for as many employees as possible. For modeling purposes, this intervention assumes that 33% of workers are able to telecommute, which is based on the reduction in travel to workplaces observed in early March, when large employers were encouraging work-from-home but before mandates against large gatherings were established.^8^
- **Cocoon seniors**. This strategy attempts to limit or delay transmission of SARS-CoV-2 to the elderly (aged ≥65 years), who are at the highest risk of severe COVID-19. In the model, this strategy assumes that nursing homes are closed persons with COVID-19 symptoms (including both visitors and staff), and that seniors in the community reduce their community contacts by 90%.
- **Community social distancing**. This intervention encourages members of the general public to reduce voluntary contacts outside of the home (not counting workplaces). The model assumes that community contacts are reduced by 25% under this intervention.
- **Self-isolation for symptoms**. Under this intervention, members of the general public are encouraged to stay home of they are experiencing symptoms of COVID-19. The model assumes that symptomatic people self-isolate 33% of the time (i.e. an average of 2.3 days per ill individual).
- **Test-and-quarantine**. This intervention aims to identify persons with SARS-CoV-2 infection, isolate the cases, and quarantine their household contacts. Cases are isolated until the end of symptoms, and household contacts are quarantined for 14 days. For modeling purposes, this intervention assumes that 20% of cases are detected on the third day of symptoms (with the other 80% never detected), and that 33% of households comply with quarantine.

To determine the potential impact of these interventions individually, each intervention was applied as of March 12^th^ 2020 (the date schools were closed) assuming that no other interventions occurred. Predicted daily hospitalizations for each individual intervention were compared to expected hospital admissions in the absence of any interventions (Figure 2).

**Figure 2:**
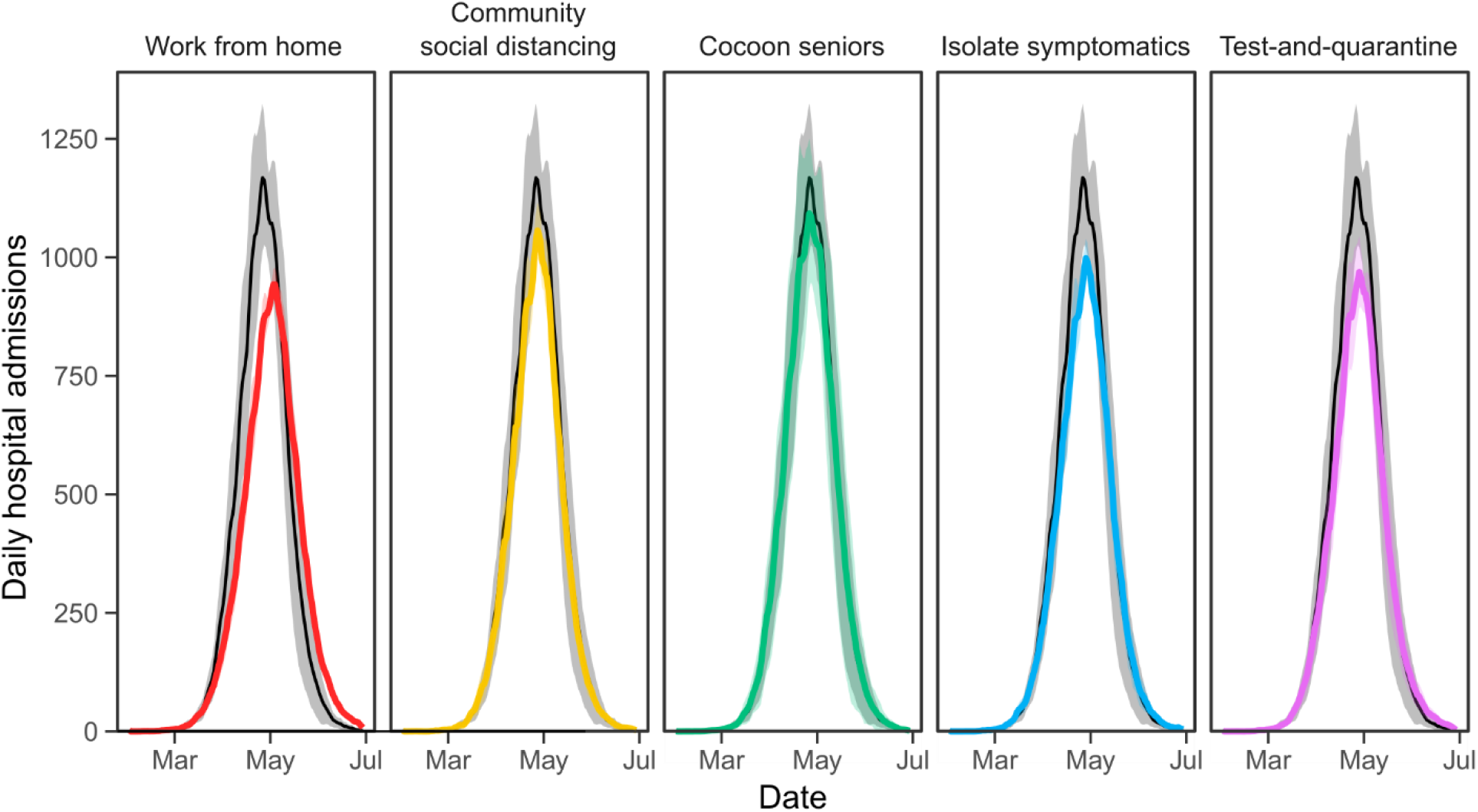
Hypothetical daily COVID-19 hospital admissions under individual mitigation strategies vs. no intervention (black lines) – King County, February – June 2020

Individually, each strategy had at most a modest impact on the expected course of the epidemic. The most effective individual strategy was test-and-quarantine, which reduced the total hospitalizations by 12.7% (95% confidence interval [CI], 12.0% to 13.3%), from a mean 42,177 hospitalizations to a mean of 36,833.

The goal of mitigation is to slow the spread of SARS-CoV-2 and protect persons at risk for severe illness, while minimizing the negative effects of intervention strategies.^9^ The aim is to identify a strategy that could allow SARS-CoV-2 to spread through the population while avoiding unnecessary deaths that could result if hospital capacity is exceeded. To this end, the number of hospital beds occupied by COVID-19 patients over time is predicted using each of the strategies tested, assuming that the interventions start on June 22^nd^ 2020. These analyses assume that the interventions replace other current or potential measures such as bans on large gatherings. Predicted hospital occupancy is compared to the King County capacity for COVID-19 hospitalizations, with a target of no more than 35% of the county’s 5,185 beds in use for COVID-19 patients.

A sequence of scenarios was tested, starting with voluntary work-from-home and adding more possible interventions. For reference, if all current COVID-19 interventions were to end on June 22^nd^, the model suggests a peak in COVID-19 hospitalizations that would exceed the available bed capacity by nearly 6-fold (Figure 3). This corresponds to a total of 41,891 (95% CI, 41,563 – 42,219) hospitalizations between June 22, 2020 and January 31, 2021. Voluntary work-from-home can reduce the overall size of the epidemic and push the peak later in the fall, but still with hospital capacity greatly exceeded.

**Figure 3:**
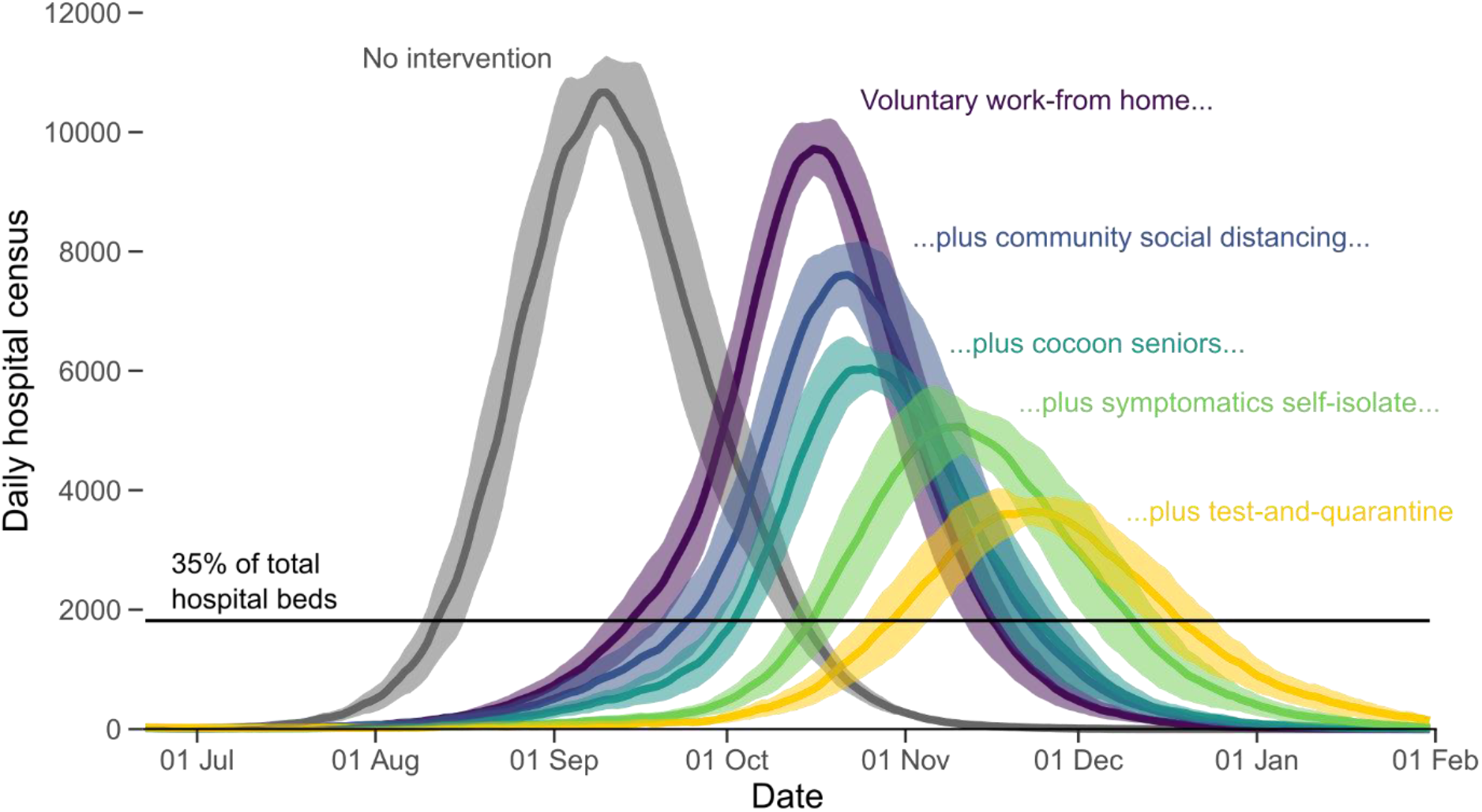
Expected number of hospital beds occupied by COVID-19 patients under cumulative intervention scenarios – King County, June 2020 – January 2021

Additional interventions further reduce the peak. All five proposed interventions together – voluntary working from home, cocooning seniors, modest social distancing, self-isolation for symptoms, and test-and-quarantine (Figure 3, yellow line) – greatly reduces the overall magnitude of the epidemic, although still in excess of the target of 35% of total hospital beds. Under this combination of interventions the effective reproductive number (R_eff_) was estimated to be 1.3 (95% CI, 1.0 – 1.6). This combination results in 20,264 (95% CI, 20,059– 20,468) hospitalizations, a reduction of 48% (95% CI, 47% - 49%) from the unmitigated prediction. The peak hospital census would occupy 70% (95% CI, 66% - 75%) of total hospital beds in King County.

Long-term school closure could further reduce COVID-19 incidence, but this is highly disruptive to students and their families. However, targeted school closures can slow transmission enough to make the epidemic manageable. As an example, Figure 4 considers a model where schools are closed for the first week of October, November, and December. This further decreases the expected number of COVID-19 hospitalizations, which remains within 35% of total hospital beds for most of the epidemic, approaching 58% of beds at the peak. Future modeling work could identify the optimal timing and duration of school closures near the peak of the COVID-19 epidemic.

**Figure 4:**
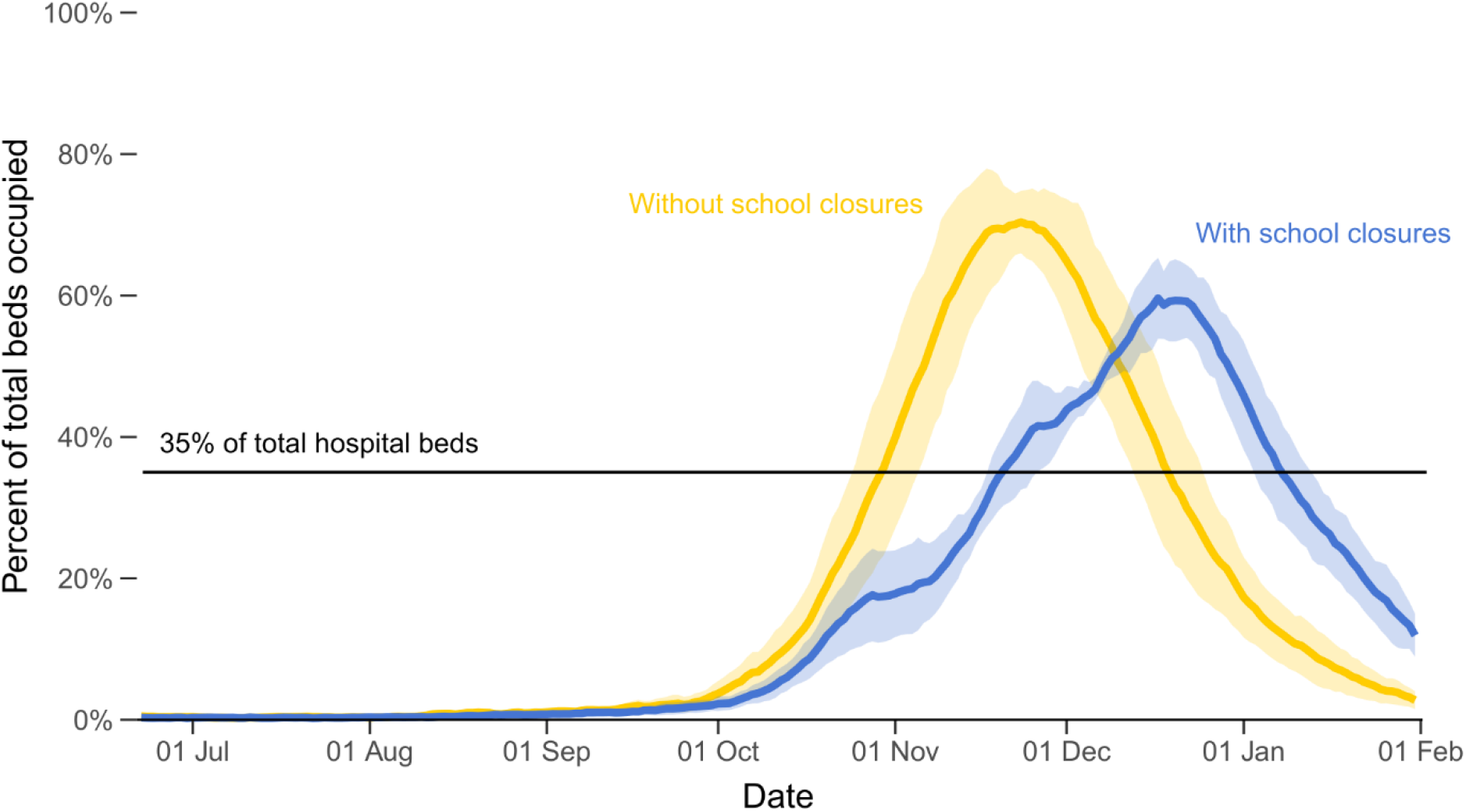
Expected proportion of hospital beds occupied by COVID-19 patients using combined interventions with or without short-term school closures – King County, June 2020 – January 2021

## Discussion

Several modeling studies have explored the potential impact of social distancing on the COVID- 19 epidemic.^10-13^ These have generally been statistical or mass-action models that explore reductions in the R_eff_ needed to achieve a certain level of control, but without identifying the actions that could create the desired R_eff_. The general conclusion of these studies is that mitigating the SARS-CoV-2 pandemic may require intermittent high-intensity interventions (reducing the basic reproductive number by 60% or so) interspersed with periods of low-intensity interventions.

Rather than estimating the impact of generic reductions in R_eff_, this report uses an agent-based model to estimate the impact of specific policies on SARS-CoV-2 transmission and COVID-19 hospitalizations. These findings suggest that layered, low-impact interventions can slow SARS-CoV-2 infection sufficiently that substantial progress can be made toward developing herd immunity while maintaining case loads at or near the capacity of the healthcare system. R_eff_ under these interventions was estimated to be 1.3, which is 48% lower than the basic reproductive number of 2.7.

Chao et al^14^ and Rosenfeld et al^15^ used also agent-based models to explore the impact of combinations of social distancing measures on SARS-CoV-2 transmission. Those analyses aimed to identify interventions to either drive R_eff_ below 1 or to maintain a low R_eff_ value. In contrast, this report focuses on strategies that allow R_eff_ to be greater than 1.0, so that there is epidemic spread of SARS-CoV-2, but ideally within the capacity of the healthcare system. The overall findings of these three reports are similar, in that combinations of measures are likely needed to make an impactful difference on SARS-CoV-2 transmission.

With the scenarios modeled here, the best set of interventions results in a peak of COVID-19 cases that would require 58% of existing hospital beds in King County. This is higher than the ideal setting, which would be keeping COVID-19 burden below the 35% of beds that are typically open. However, this level of usage could still be met within the system through additional actions during the peak of incidence such as postponing elective surgeries. Future modeling work could also explore additional strategies that may further “flatten the curve” to keep hospitalizations within ideal levels.

This agent-based model makes several key assumptions which may impact its accuracy. First, for computational simplicity, the model does not include demographic processes – births, deaths, or aging. Over short model runs (≤1 year) this assumption is unlikely to meaningfully impact model forecasts, but the lack of births (which introduce new Susceptible individuals) would become unrealistic over longer runs. Second, the model assumes that all Infectious individuals are equally infectious. The model includes the possibility of superspreading *events*, where a single individual could infect many contacts by chance, but does not including superspreading *agents*, who are inherently more likely to cause secondary infections. Third, the model assumes that Infectious agents are equally infectious during their entire infectious period. Fourth, while the probability of severe disease among infectious agents varies by age, the probability of infection given contact with an infectious agent is assumed to be identical for all susceptible agents. Fifth, the model assumes that SARS-CoV-2 transmission does not vary meaningfully by season. Finally, the model was developed based on the demographics of King County Washington, and findings may not generalize to other areas with different population densities or age structures.

Individual low-impact interventions may not substantially reduce the transmission of SARS-CoV-2. However, combining several interventions may be able to mitigate the course of the COVID- 19 epidemic in King County by keeping hospital burden within or near the capacity of the healthcare system. Under this approach the virus can spread through the community, moving toward herd immunity, while minimizing the social and economic disruption of the control measures.

## Methods

The simulation model is implemented using the GAMA platform.^16^ The model code and data are publicly available at http://github.com/ThatFluGuy/nCoV_ABM.

### Model population

The model simulates a population of individual people, known as “agents,” living in a large metropolitan area. Agents in the model are divided into households such that the demographic (age and sex) distribution of the agents reflects the King County household composition, according to the 2018 American Community Survey.^6^ The model also contains locations, which include households, schools, workplaces, community gathering sites (e.g. churches, fitness centers, restaurants), and nursing homes and other group quarters.

While the full population of King County is approximately 2.23 million persons, the simulation includes approximately 509,000 agents to make the model computationally feasible. To facilitate comparisons with observed data, estimated hospitalization counts from the model output are scaled up by a factor of 4.38 to match the size of the actual King County population.

Agents are divided into different classes, which define the types of locations they can travel to during the simulation. Examples include children aged <6 years who do not attend daycare or pre-kindergarten outside the home; school-aged children who go to school during the work week; working adults; community-dwelling seniors; or nursing home residents.

The model is stochastic, in that events such as transmission of infection have a pre-determined probability of occurring but occur by random chance in an individual run of the model. All simulation data presented are generated by running multiple iterations of the model and averaging the results.

### Time and movement

The simulation moves through discrete time steps, with each day divided into three segments: workday, afternoon, and evening. During the workday on weekdays, children go to school and adults with a workplace go to that workplace. Schools, nursing homes, and group quarters are workplaces for some adults. The number of schools is selected assuming that children at school make effective contact (i.e. contact sufficient to transmit SARS-CoV-2 from an infectious to a susceptible agent) with up to 100 other children at school. Workplace sizes are distributed according to the business sizes from the 2018 Small Business Profile for Washington State,^17^ with the assumption that workers in businesses of more than 100 persons only make effective contact with up to 100 workers per day.

During the afternoon, all community-dwelling agents may visit a random community site, with greater probability on weekends than weekdays. In the evening, all agents go to their household. Nursing home and group quarter residents do not move during any time step.

### Observed interventions

A series of progressive interventions was implemented in Washington State to suppress the escalating SARS-CoV-2 epidemic. For King County, this included prohibiting large gatherings (March 11^th^); closing schools (March 12^th^); closing restaurants, bars, entertainment facilities, and prohibiting mid-size gatherings (March 16^th^); and finally shelter-in-place orders on March 23^rd^. These official social-distancing restrictions were accompanied by voluntary actions by businesses and private individuals, such as multiple large employers encouraging employees to work from home as of March 2^nd^.

Data from Facebook’s “Data for Good Project”, as compiled by the Institute for Disease Modeling,^8^ were used to estimate the percent reduction in out-of-home travel during different intervention stages. Schools in the model were closed as of March 12^th^, so that children and teachers were assumed to stay home during the workday.

### SARS-CoV-2 transmission

With respect to SARS-CoV-2, each agent is characterized as one of four mutually exclusive categories: Susceptible, Exposed (infected but not yet infectious), Infectious, and Recovered (and immune). When a Susceptible agent shares a location with one or more Infectious agents, the Susceptible agent may become infected. The probability of infection for each Infectious contact is higher within households than in the community.^18,19^ Infected agents move into the Exposed category and are randomly assigned a latent period (until onset of infectiousness) and an incubation period (until onset of symptoms) (Table). The latent period is between 3 and 5 days, and the incubation period is assumed to be 1-3 days longer than the latent period,^20^ so that individuals become infectious before becoming symptomatic. At the end of the latent period, agents become Infectious and are randomly assigned an infectious period of 6-8 days. Agents are assumed to be equally infectious regardless of age or time since infection. Infectious individuals have an age-specific probability of being hospitalized; probabilities range from 0.1% among agents aged <1 year to 30% among those aged 99 years, based on age-specific incidence of COVID-19 hospitalization in Washington and California.^21^ Hospitalized agents enter the hospital 3-5 days after onset of symptoms.^22^ Transmission in hospital settings is ignored. At the end of the infectious period, agents become Recovered and are assumed to be immune to infection for the duration of the simulation.

**Table:** Values for SEIR model parameters

| Model parameter | Values | References |
| --- | --- | --- |
| Latent period (days) | 3-5 days | Li et al,<sup>24</sup> Peirlinck et al<sup>25</sup> |
| Incubation period (days) | Latent period + 1-3 days | Li et al,<sup>26</sup> Lauer et al,<sup>27</sup> Linton et al<sup>22</sup> |
| Infectious period (days) | 6-8 days | He et al<sup>20</sup> |
| Duration of hospitalization (days) | 11 days | Guan et al,<sup>28</sup> Docherty et al,<sup>29</sup> Shah et al<sup>30</sup> |
| Per-contact probability of infection (households) | 0.024 | Estimated from model fitting |
| Per-contact probability of infection (non-household locations) | 0.008 | Estimated from model fitting |

The per-contact risks of infection in households and communities were estimated from cumulative daily hospital admissions data for King County,^23^ with the observed interventions as described above. A scaling factor was estimated to translate Facebook Data for Good travel frequencies to reduced workplace and community site visit frequencies. Both the scaling factor and infection risks were estimated by fitting simulated cumulative daily hospital admissions to observed counts using a tabu algorithm. The range of possible values for per-contact infection risks was selected to allow a basic reproductive number (Ro) in the range of 2 to 5. After fitting the parameters, Ro was estimated by running 200 iterations of the simulation, starting with one random agent infectious, and counting the number of secondary infections from that agent.

## Data Availability

Data and code are available http://github.com/ThatFluGuy/nCoV_ABM

